# Symptom network signatures for the early recognition of pancreatic cancer

**DOI:** 10.64898/2026.02.22.26346814

**Authors:** Jaymart G. Latigay, Louie F. Dy, Geoffrey A. Solano

**Affiliations:** College of Arts and Sciences, University of the Philippines Manila, Manila, Metro Manila, Philippines; Office of the Deputy Director for Health Operations, University of the Philippines-Philippine General Hospital, Manila, Metro Manila, Philippines

## Abstract

**Background:** Pancreatic cancer is a leading cause of cancer mortality, and early recognition is challenging. To achieve early diagnosis using symptoms alone, we examined patterns across different stages using network analysis to derive clinically useful insights.

**Methods:** Symptom variables from a de-identified dataset of 50,000 pancreatic cancer patients were analyzed. Stratification by stage was done, followed by bootstrap resampling to address imbalances across strata. Symptom networks were then constructed with nodes representing symptoms and edges representing conditional dependencies estimated via an Ising-style neighborhood selection approach implemented through L1-regularized logistic regression. Strength, betweenness, and closeness centrality indices were then calculated, and their stability was analyzed using the case-dropping bootstrap. Network comparison tests were done, and difference networks were analyzed. Spring-layout algorithm was used for visualization, with node size being the predictability (pseudo-R²), and the edge weight being the mean partial correlation magnitude.

**Results:** On average, symptoms were present in about one out of four patients (M = 0.26). Weight loss and abdominal discomfort were the most prevalent of the symptoms, followed by jaundice and back pain. Network structures became sparser across stages with a decreasing number of edges and centrality indices. Jaundice emerged as the dominant hub in Stage I, but shared dominance with Weight Loss in Stage II. Node predictability (pseudo-R^2^) was effectively zero across all disease stages.

**Conclusion:** Our network analysis of pancreatic cancer symptomatology across stages revealed distinct patterns that may improve understanding of its clinical presentation and support earlier recognition.

## INTRODUCTION

Pancreatic cancer is the seventh leading cause of cancer mortality in the world^[1]^. Most present with advanced or metastatic disease (Stages III-IV), and five-year survival is poor: 10% for all stages, 3% for metastatic disease, 15% for regional disease, and 36% for localized disease^[1]^. In the Philippines, the average life expectancy is less than one year, with a five-year survival rate of 2%, making it the eighth leading cause of cancer death^[2]^. There is no proven, reliable strategy for achieving early-stage diagnosis using symptoms alone in the general population. There are also currently no biomarkers with sufficient sensitivity and specificity. Currently, screening is done primarily in high-risk individuals^[3]^. Some of the risk factors include hereditary cancer syndromes such as Peutz-Jeghers syndrome or familial history of pancreatic cancer, age, male sex, smoking, heavy alcohol drinking, red meat consumption, chronic pancreatitis, diabetes, and obesity^[3]^. While novel approaches, such as liquid biopsies, artificial intelligence, metabolomics, ion mobility spectrometry (IMS) associated technologies, and novel nanomaterials have shown good potential for clinical translation^[3]^, these are not immediately available, especially in resource-constrained, primary care settings. This study aims to examine patterns in symptoms of pancreatic cancer across different stages using network analysis and identify signatures that may aid in the early recognition of pancreatic cancer for both clinicians and laypersons.

## METHODS

### Data Acquisition and Preparation

This study utilized a de-identified, open-access dataset of approximately 50,000 pancreatic cancer patients, sourced from several undisclosed international institutions between April 20, 2021, and July 21, 2024. The dataset is publicly hosted on the Kaggle repository under the title “Pancreatic Cancer Prediction Dataset,” published by Ankush Panday on the platform^[4]^. The dataset contained 24 variables, including patient demographic information, risk factors, and commonly reported symptoms. Data quality assessment revealed minimal data issues consisting of a limited number of duplicates and incomplete entries. These were subsequently removed, given that their impact on the overall sample size was negligible. Verification of binary representation (0 = absence, 1 = presence) was also performed to ensure consistency.

To focus the analysis only on clinical presentation and symptom profiles, the dataset was filtered to include only the stage of diagnosis (I to IV) and four specific symptoms: “Jaundice,” “Weight Loss,” “Abdominal Discomfort,” and “Back Pain.” Demographic and risk factor variables were excluded, as these were already well-established in the medical literature^[1]^. All data management and analysis were conducted in Python 3.10 using pandas, NumPy, scikit-learn, NetworkX, and Matplotlib as the main libraries.

### Statistical Analysis

Descriptive statistics were computed to summarize the study population. Categorical variables and binary symptoms were expressed as frequencies and percentages. To characterize symptom distribution across disease progression, the prevalence of Jaundice, Weight Loss, Abdominal Discomfort, and Back Pain was calculated and stratified by cancer stage (Stages I to IV).

### Network Estimation and Bootstrap Stability

Network analyses were stratified by the stage at diagnosis to identify stage-specific symptom interactions and observe how co-occurrence patterns evolved with disease progression. In these networks, nodes represented individual symptoms, and edges denoted conditional dependencies between two symptoms after controlling for all others. The network structure was estimated using an Ising model framework via L1-regularized (Lasso) logistic regression, which identifies sparse, interpretable connections among binary variables^[5], [6]^. Specifically, each symptom was regressed on all other symptoms using logistic regression with L1 penalty, and the regularization parameter was selected via 4-fold cross-validation across a logarithmic grid ranging from 0.001 to 1000. Edge weights were symmetrized using the AND-rule, which takes the maximum absolute coefficient from both regression directions to produce undirected edges representing symmetric conditional associations.

Although the sample size varied across stages (Stage I: 4,937; Stage II: 10,173; Stage III: 14,968; Stage IV: 19,918), the dataset provided sufficient statistical power for all the groups due to their individual sizes. To ensure that the identified associations were robust regardless of sample size, a non-parametric bootstrap procedure was integrated into the estimation process^[7]^. For each stage, 200 bootstrap iterations were performed with 75% resampling. Edges were only retained if they appeared in at least 60% of the bootstrap samples (frequency threshold ≥ 0.60). This ensured that the final networks generated reflected a stable, high-confidence symptom interaction for every stage.

### Bootstrap Confidence Intervals and Consistency

To quantify the uncertainty of edge weight estimates, 95% confidence intervals were calculated for all stable edges using the percentile method across bootstrap distributions^[7]^. Additionally, bootstrap consistency was assessed by splitting the 200 bootstrap samples into two equal halves and computing the correlation between edge weight estimates from each half. High correlation values (> 0.7) indicated consistent and reliable edge weight estimation across different data subsets.

### Network Measures

To characterize the structural importance of each symptom, three centrality indices were computed for the stage-specific networks: Strength, Betweenness, and Closeness^[8]^. Strength Centrality was calculated as the sum of the absolute edge weights connected to a node, reflecting the overall magnitude of associations. Betweenness Centrality quantified the frequency with which a node lies on the shortest path between other nodes, identifying potential “bridge” symptoms. Closeness Centrality was measured as the inverse of the average shortest path length from a symptom to all others, indicating how quickly a symptom interacts with the rest of the network.

Additionally, node predictability was computed to assess how well each symptom could be explained by its neighbors in the network. This was quantified using pseudo-R^2^ coefficient derived from logistic regression models, where each symptom was predicted by all other symptoms^[9]^. Higher predictability values indicate that a symptom’s presence is strongly determined by network structure, whereas lower values suggest independence.

### Centrality Stability Analysis

To assess the reliability of node importance rankings, centrality stability coefficients (CS-coefficients) were computed using a case-dropping bootstrap^[7]^. This procedure tests whether centrality rankings remain consistent when a proportion of cases is randomly removed from the dataset. For each stage and centrality measure (strength, betweenness, and closeness), 100 bootstrap samples were generated with 10%, 25% 50% and 75% of cases randomly dropped. For each subsample, the network was re-estimated, and centrality values were recalculated. Spearman rank correlations between full-sample centrality rankings and subsample rankings were computed, with the CS-coefficient defined as the mean correlation across all drop percentages. Following established guidelines^[7]^, CS-coefficients > 0.7 indicate excellent stability, > 0.5 indicate acceptable stability, > 0.25 suggest interpretability with caution, and < 0.25 indicate unstable rankings that should not be interpreted.

### Network Comparison Tests

To formally test whether networks differed significantly between consecutive cancer stages, Network Comparison Tests (NCT) were performed with 1,000 iterations^[5]^. Group labels (stage membership) were randomly shuffled, networks were re-estimated for each permuted dataset, and the distribution of test statistics under the null hypothesis of no difference was generated.

01Three complementary tets were conducted: (1) overall network structure test, using the maximum absolute edge difference as the test statistic to assess global structural differences; (2) global strength test, using the absolute difference in the sum of all edge weights to test overall connectivity changes; and (3) edge-specific tests, computing individual edge differences and performing two-tailed tests to identify which specific symptom associations changed significantly between stages^[5]^. Statistical significance was defined as p < 0.05.

### Difference Networks

To visualize stage-to-stage changes in symptom relationships, difference networks were constructed by subtracting the adjacency matrices of consecutive stages (e.g., Stage II - Stage I)^[5]^. Only stable edges (frequency ≥ 0.60) in at least one of the two networks being compared were retained in the difference network. Positive values indicated edges that were stronger in the later stage, while negative values indicated edges that were stronger in the earlier stage. Edge thickness in visualization was proportional to the magnitude of change.

### Network Visualization

Symptom networks per cancer stage were visualized using Fruchterman-Reingold spring-layout algorithm^[10], [11]^ via NetworkX and Matplotlib. In these visualizations, nodes with stronger connections are positioned closer together through force-directed placement, where edges act as springs that attract connected nodes, while nodes repel each other. Edge width corresponds to the strength of association (mean partial correlation weight from bootstrap samples), while node size reflects node predictability (pseudo-R^2^). In stage-specific network visualizations, all edges are displayed in green. In different networks, edge coloring distinguishes the direction of change: blue edges represent associations that are stronger in the later stage, while red edges represent associations that are stronger in the earlier stage.

## RESULTS

### Descriptive Statistics

Mean, standard deviation, skewness, kurtosis, and frequency (absence and presence) of the clinical symptoms were summarized in Supplementary Table 1 for each stage of pancreatic cancer. Across all stages, the average symptom mean was approximately M = 0.26 (SD = 0.4), indicating that, on average, symptoms were present in about one out of four patients. Weight loss and abdominal discomfort consistently showed the highest mean values across all stages, suggesting these were the most commonly reported symptoms among individuals diagnosed with pancreatic cancer. On the other hand, jaundice and back pain were slightly less prevalent, with mean presence rates ranging between 0.19 and 0.25.

Across stages, the distribution of symptom presence was positively skewed, reflecting that most patients did not report these symptoms. Skewness values ranged from 0.62 to 1.54, while kurtosis values ranged from 1.62 to 0.36, indicating light-tailed, right-skewed distributions typical for binary symptoms data. The standard deviation was consistently higher for weight loss and abdominal discomfort (SD ≈ 0.47), suggesting slightly greater variability in these symptoms across patients compared to jaundice and back pain.

Symptom frequency patterns were largely stable across disease progression. Minor, negligible increases were observed in each of the symptoms, around 1%, from stage I to IV. This suggested that, at least in this dataset, the overall pattern of symptom occurrence remained consistent across stages.

### Network Structure and Complexity

The symptom networks demonstrated progressive simplification (decrease in the number of edges) with advancing disease stage (Table 1, Figure 1). Stage I networks contained 5 stable symptom associations (network density = 0.719), reflecting the highest complexity of symptom co-occurrence patterns amongst the cancer stages. This declined substantially through intermediate stages: Stage II contained 4 stable edges (network density = 0.493), depicting a 31% reduction in network density from Stage I. However, the most dramatic simplification occurred between Stage II and III, where networks consolidated to only 2 stable edges (network density = 0.362), a 26% reduction. The Stage IV networks remained sparse with only 2 stable edges (density = 0.318), indicating a plateau of network simplification at advanced stages of the disease.

**Figure 1.**
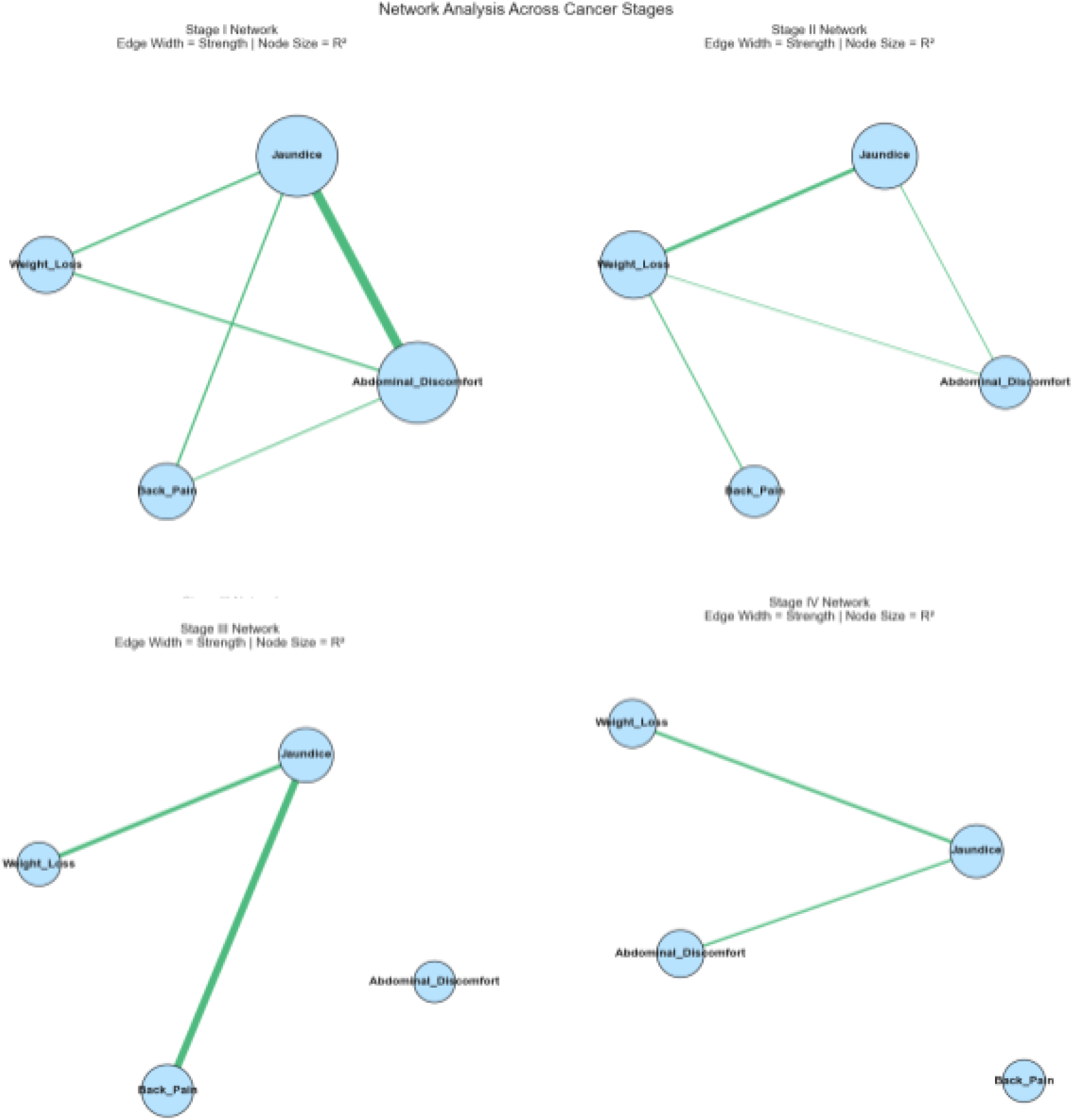
Symptom networks across pancreatic cancer stages

**Figure 2.**
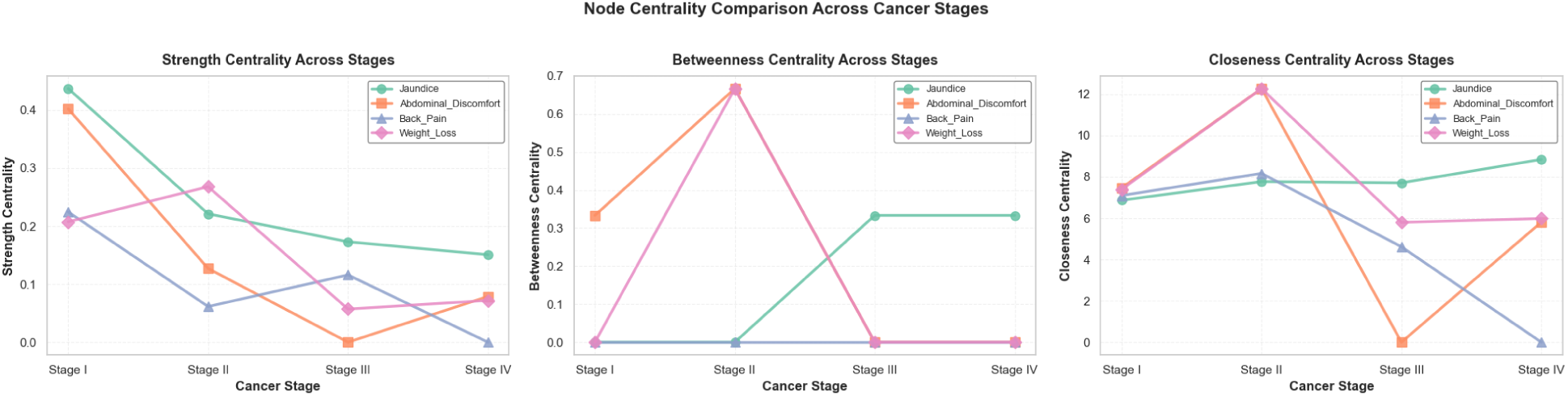
Node centrality comparison across pancreatic cancer stages

**Figure 3.**
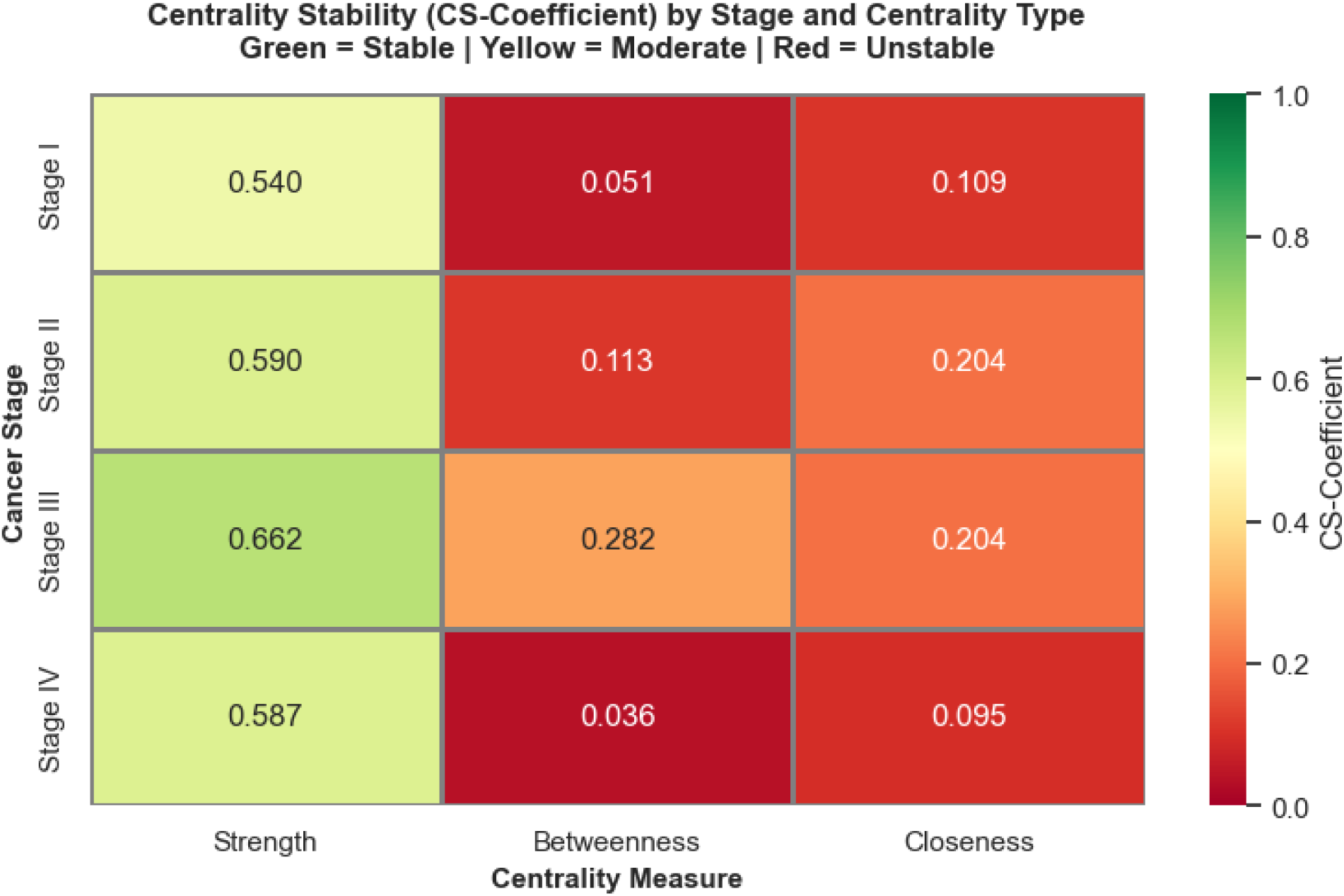
Centrality stability by stage and centrality type

**Figure 4.**
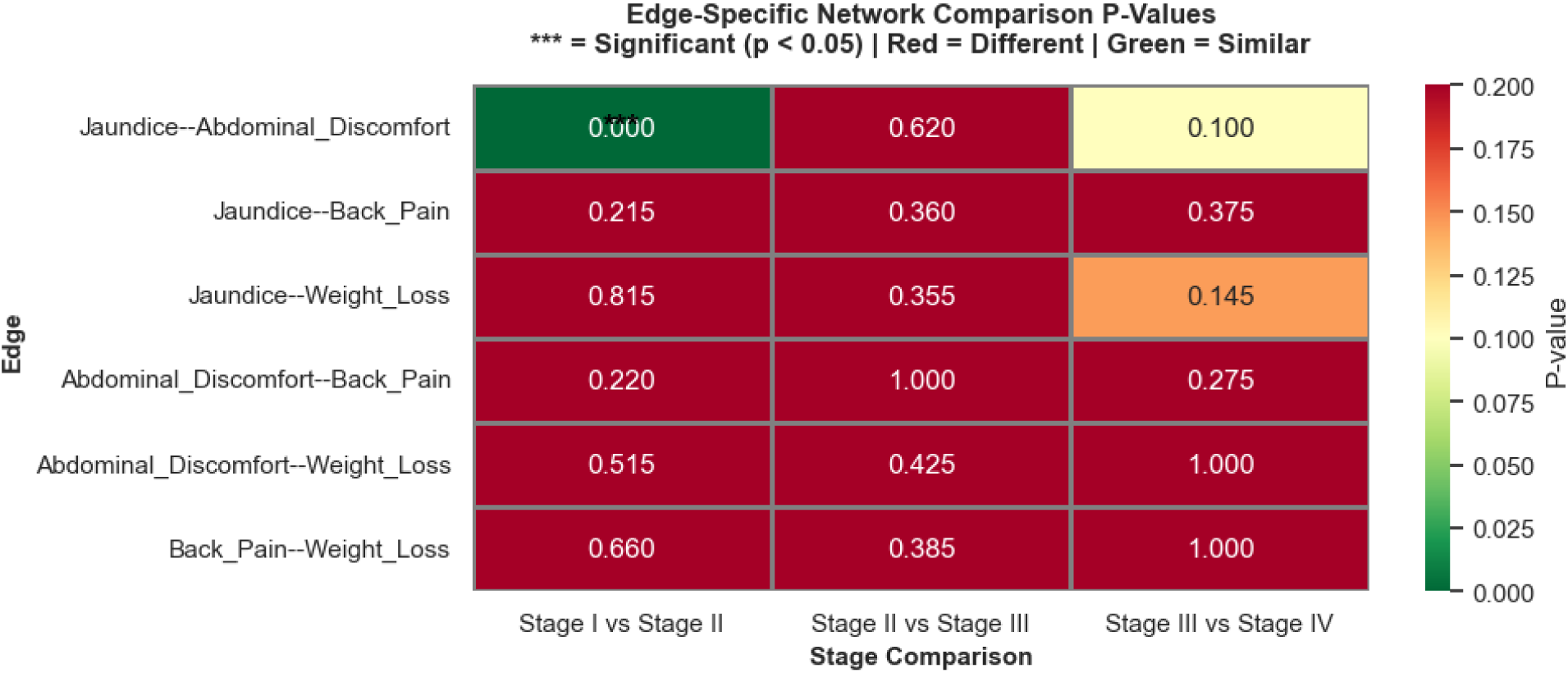
Edge-specific network comparison of p-values

**Figure 5.**
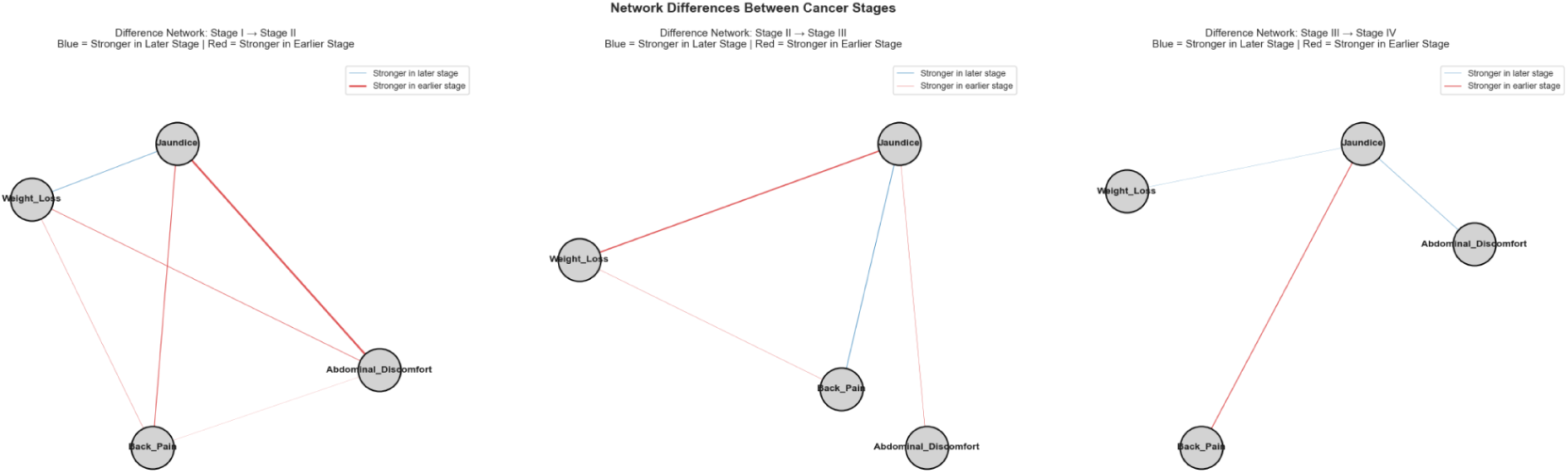
Network difference between pancreatic cancer stages

**Table 1.**
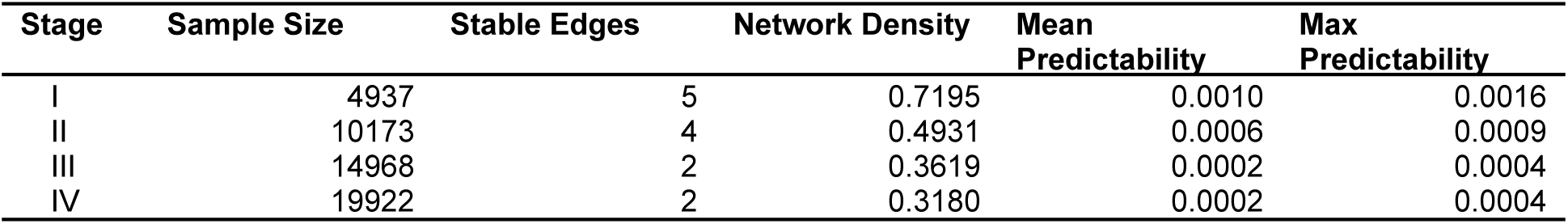
Summary of network metrics by stage.

| Stage | Sample Size | Stable Edges | Network Density | Mean Predictability | Max Predictability |
| --- | --- | --- | --- | --- | --- |
| I | 4937 | 5 | 0.7195 | 0.0010 | 0.0016 |
| II | 10173 | 4 | 0.4931 | 0.0006 | 0.0009 |
| III | 14968 | 2 | 0.3619 | 0.0002 | 0.0004 |
| IV | 19922 | 2 | 0.3180 | 0.0002 | 0.0004 |

Bootstrap stability assessment revealed important limitations in edge weight precision. Cross-validation of edge weights between the first and second halves of bootstrap samples (n = 200 iterations) yielded correlation coefficients close to 0 across all stages (Stage I: r = 0.0014; Stage II: r = 0.0407; Stage III: r = −0.0021; Stage IV: r = 0.0026), indicating substantial variability in edge weight estimates across bootstrap replicates. In addition, the 95% confidence intervals for stable edges were notably wide, ranging from 0.11181 to 0.3985 across all stages (mean width = 0.213), indicating considerable uncertainty in point estimates of edge weights. These findings suggested that while edge presence (frequency ≥ 60%) was relatively stable and reproducible, the precise magnitude of edge weights varied considerably depending on which data subset was sampled. Consequently, network topology can be interpreted with reasonable confidence, but the quantitative comparison of the edge weights between stages should be interpreted cautiously due to their wide confidence bounds and low cross-sample correlations.

### Symptom Centrality Patterns

#### A. Strength / Weighted Degree Centrality (Association Strength)

Jaundice emerged as the dominant network hub in Stage I with a strength centrality of 0.437, slightly higher than that of Abdominal Discomfort with 0.402 and nearly twofold higher than Back pain with 0.225. During the transition to Stage II, Weight Loss substantially increased in importance, becoming co-dominant with strength centrality of 0.268, while Jaundice and Abdominal Discomfort declined more than two-fold with 0.221 and 0.127. In advanced stages (III and IV), Jaundice reasserted its dominance amongst the symptoms but at substantially reduced absolute values (Stage III: 0.173; Stage IV: 0.151), reflecting the overall network sparsification at these stages.

#### B. Betweeness Centrality (Bridging Roles)

Betweenness centrality revealed stage-specific patterns of symptom bridging. In Stage I, only abdominal discomfort displayed bridging capabilities with a score of 0.3333, while the rest had 0. In the succeeding stage, Stage II, both Weight Loss and Abdominal Discomfort demonstrated co-dominant bridging activity with betweenness centrality of 0.667, substantially higher than Jaundice and Back Pain, which both had 0. This bridging role shifted in advanced stages, in stage III and IV, wherein Jaundice emerged as the primary bridge with 0.333 in both stages, while Weight Loss and Abdominal Discomfort lost their bridging function. These patterns indicate a stage-dependent reorganization of network topology, with different symptoms assuming mediating roles as cancer progresses.

#### C. Closeness Centrality (Network Accessibility)

In Stage II, Weight Loss and Abdominal Discomfort both showed exceptionally high closeness, with both scores being 12.27, indicating the most efficient access to all other symptoms. Stage I closeness values were moderate (6.87 to 7.46), and those of Stage III declined further (0 to 7.71), with Abdominal Discomfort getting a 0 closeness score. In Stage IV, only Back Pain collapsed to 0, while Jaundice (8.84), Weight Loss (5.99), and Abdominal Discomfort (5.81) retained moderate accessibility.

### Centrality Stability Assessment

Strength centrality exhibited acceptable stability across all stages. Case-dropping bootstrap analysis (10 to 75% removal with 100 iterations per level) produced CS-coefficient of 0.540, 0.590, 0.662, and 0.587 for Stages I to IV, respectively. Strength-based rankings remained reasonably reliable under case dropping, given that all exceeded the 0.5 threshold.

Betweenness stability was low in most stages, with scores ranging from 0.036 to 0.113, but reached a cautionary level in Stage III (0.282), indicating that bridging rankings were generally unreliable and only tentatively interpretable in Stage III. Closeness stability was consistently unstable, with scores from 0.095 to 0.204 across all stages, suggesting that closeness-based ranking should not be used for interpretation.

### Node Predictability

Node predictability (pseudo-R^2^) was effectively zero across all stages, indicating that symptom presence was almost entirely unexplained by network neighbors under the current model. Mean (Max) pseudo-R² values were Stage I: 0.0010 (0.0016); Stage II: 0.0006 (0.0009); Stage III: 0.0002 (0.0004); Stage IV: 0.0002 (0.0004). There was no meaningful stage-to-stage decline, as predictability was already near zero in the early stages and remained close to zero in advanced stages. These results suggested that, in this four-symptom specification and logistic regression modeling setup, symptom occurrences behave largely independently with respect to variance explained by the estimated network.

### Stage-to-Stage Network Transitions

#### A. Network Comparison Results

Permutation testing revealed that symptom networks remained largely stable throughout disease progression. Between Stage I and Stage II, networks showed no significant differences in overall structure (maximum edge difference = 0.176, p = 0100) or global connectivity strength (maximum edge difference = 0.464, p = 0.315), with only one edge showing statistical significance with a p-value < 0.0001. The Stage II-to-Stage III transition similarly showed no significant structural differences (maximum edge difference = 0.093, p = 0.620; strength difference = 0.126, p = 0.760), with no individual edge reaching statistical significance. The Stage III-to-Stage IV transition also showed no significant differences (p = 0.455 for structure; p = 0.590 for strength), indicating network continuity at advanced disease stages.

#### B. Edge-Specific Changes

Individual edge analysis revealed differential robustness of symptom associations. The Jaundice-Abdominal Discomfort edge differed significantly between Stage I and II (p < 0.0001), The Jaundice-Weight Loss edge remained remarkably stable across all stage transitions (p ≥ 0.145), establishing this as a fundamental association throughout disease progression. The Jaundice-Back Pain and Abdominal Discomfort-Back Pain edges showed borderline non-significance at Stage I-to-Stage II transition (p = 0.215 and 0.220, respectively), while remaining stable at later transitions. The Abdominal Discomfort-Weight Loss and Back Pain-Weight Loss edges showed consistent stability across all stage transitions (all p ≥ 0.385).

### Temporal Network Evolution

The Stage I-to-Stage II transition featured increased strength of the Weight Loss-Jaundice association (+0.041), indicating modest tightening of these symptoms in the intermediate stage. The most substantial change occurred within the Stage II-to-Stage III transition, wherein the Weight Loss-Jaundice association weakened (−0.093), representing a 35% reduction in this edge weight, while the Jaundice-Back Pain association strengthened (+0.051). The Stage III-to-Stage IV transition showed that while some changes persisted (Jaundice-Back Pain association weakened by −0.066), the overall magnitude of differences decreased compared to earlier transitions.

## DISCUSSION

### Summary of Major Findings

Our study examined patterns in symptoms of pancreatic cancer across different stages using network analysis to derive clinically useful insights that may aid in the early diagnosis of pancreatic cancer. We found that symptoms were present in about one out of four patients. Weight loss and abdominal discomfort were the most prevalent of the symptoms, followed by jaundice and back pain.

There was a consistent pattern of progressive symptom network simplification concurrent with advancing pancreatic cancer stage. Early-stage networks exhibited greater complexity and higher edge density (Stage I density: 0.719), with complex multi-symptom interactions reflecting diverse clinical presentations. Network structure became progressively simplified through Stage III (Stage III density: 0.362), followed by stabilization in Stage IV (Stage IV density: 0.318)

Symptom importance, as measured by strength centrality, showed dynamic reorganization across stages. Jaundice emerged as the dominant hub in Stage I, but shared dominance with Weight Loss in Stage II. This shifting centrality hierarchy, combined with high CS-coefficient stability (≥ 0.540 across all stages), establishes a strength-based measure as a reliable indicator of symptom importance.

Node predictability (pseudo-R^2^) was effectively zero across all disease stages, indicating that symptom occurrence was almost entirely unexplained by network neighbors under the current modeling framework. Symptoms may have stable connections within the network, but do not necessarily predict each other (i.e., are mostly conditionally independent).

### Clinical Correlation

The symptom network structures were consistent with the known underlying disease progression of pancreatic cancer. Stage I showed the strongest connections between jaundice and abdominal discomfort, analogous to localized (confined) disease where pancreatic head masses compressed the common bile duct, leading to biliary obstruction and peritoneal irritation with more pronounced symptoms. Stage II showed weight loss as the central hub, yet still maintaining moderately-strong connections with jaundice, abdominal discomfort, and back pain, implying local disease with beginning systemic manifestations, as tumor cells spread to nearby lymph nodes but not distant sites. Stage III and IV disease showed a sparse network, suggesting variability, inconsistency, and conditional independence of symptoms. We found this consistent with the clinical Stage III, defined by nodal metastases and invasion into nearby major blood vessels, and clinical Stage IV, defined by metastases to distant organs. Moreover, pancreatic tail tumors are easily clinically silent during the early stages, which may not even be noticed by either physicians or patients for several months to years, until they metastasize into the liver or spine, which *then* leads to symptoms such as jaundice, abdominal discomfort, and back pain.

Because edges signified conditional dependence, each symptom network may be read as “symptom X may be used to inform symptom Y”, such as, in Stage II, weight loss may be used to inform jaundice and abdominal discomfort. This also suggests a clinical application, such as when doctors interview and ask patients about their symptoms.

Weight loss, while clinically may be only of minor consideration (or even be seen as a sign of being healthy and “fit”) for both clinicians and patients, emerged as the central hub that serves as an early sentinel for the latter stages (advanced - Stage III or metastatic - Stage IV) of pancreatic cancer in our network analysis. Jaundice remained the most accessible symptom as it had the highest closeness centrality in Stage I and III. This is consistent with clinical practice, where the presence of jaundice, alongside abdominal pain, raises the suspicion of hepatobiliary diseases such as gallstones, hepatitis, and pancreatic cancer.

### Limitations of the Study

Although descriptive network metrics revealed substantial structural differences between stages (e.g., progressive density reduction, changing edge patterns), quantitative permutation testing did not detect statistically significant differences in global network structure or connectivity strength between any adjacent stage pairs (all > 0.05). This discrepancy between descriptive patterns and inferential statistics may reflect high within-stage variability or limited statistical power for network-level comparisons. The lack of statistical significance suggested that stage-to-stage network changes, while descriptively apparent, should be interpreted cautiously.

Heterogeneity among cancer patients was expected as the dataset was sourced from several undisclosed international institutions over three years, and the actual data collection methods were unclear and of high risk of bias (i.e., likely to be largely retrospective). The dataset may be fraught with recording or recall bias, and it also did not record the temporal sequencing (order) of symptoms.

Moreover, pancreatic cancer, especially in the advanced stages, would definitely have more symptoms arising from complications, such as, but not limited to, cough or shortness of breath (from lung metastases), leg pain or cramps (deep venous thrombosis), hematemesis (gastrointestinal bleeding, tumor rupture), headache or altered sensorium (from brain metastases), among many others. These were not captured by our data source, limiting our interpretation of the network structures in the advanced stages.

## CONCLUSION

Our network analysis of pancreatic cancer symptomatology across stages revealed distinct symptom structures and signatures at each stage, which may improve understanding of its clinical presentation and support earlier recognition.

## RECOMMENDATIONS

This approach might be helpful for many other diseases, not just cancers, with vague prodromal symptoms. For future studies, we recommend validation studies of symptom networks in larger, prospective cohorts that account for duration and order of a greater variety of symptoms. We also suggest looking into other metrics and analysis methods, such as accuracy metrics (sensitivity, specificity, AUROC) and complexity metrics (entropy). We recommend spreading awareness, designing cancer screening programs, and conducting further epidemiological research among patients who present with clinically significant, unintentional weight loss.

## Data Availability

All data produced in the present work are contained in the manuscript.

https://doi.org/10.34740/KAGGLE/DSV/10627162

## APPENDICES

**Supplementary Table 1.** Mean, standard deviation, minimum, maximum, skewness, kurtosis, and frequency of the CDI Symptoms by stage.

| **Stage I (n = 4937)** |  |  |  |  |  |  |  |  |
| --- | --- | --- | --- | --- | --- | --- | --- | --- |
|  | Mean | Std | Min | Max | Skewness | Kurtosis | Absence % | Presence % |
| Jaundice | 0.195463 | 0.396597 | 0 | 1 | 1.535908 | 0.359013 | 80.45372 | 19.54628 |
| Abdominal_D<br>iscomfort | 0.284181 | 0.451069 | 0 | 1 | 0.957021 | -1.08411 | 71.58193 | 28.41807 |
| Back_Pain | 0.254608 | 0.435685 | 0 | 1 | 1.12658 | -0.73082 | 74.53919 | 25.46081 |
| Weight_Loss | 0.352643 | 0.477841 | 0 | 1 | 0.616823 | -1.61953 | 64.73567 | 35.26433 |

| **Stage II (n = 10173)** |  |  |  |  |  |  |  |  |
| --- | --- | --- | --- | --- | --- | --- | --- | --- |
|  | Mean | Std | Min | Max | Skewness | Kurtosis | Absence % | Presence % |
| Jaundice | 0.195463 | 0.396597 | 0 | 1 | 1.535908 | 0.359013 | 80.45372 | 19.54628 |
| Abdominal_D<br>iscomfort | 0.284181 | 0.451069 | 0 | 1 | 0.957021 | -1.08411 | 71.58193 | 28.41807 |
| Back_Pain | 0.254608 | 0.435685 | 0 | 1 | 1.12658 | -0.73082 | 74.53919 | 25.46081 |
| Weight_Loss | 0.352643 | 0.477841 | 0 | 1 | 0.616823 | -1.61953 | 64.73567 | 35.26433 |

| **Stage III (n = 14968)** |  |  |  |  |  |  |  |  |
| --- | --- | --- | --- | --- | --- | --- | --- | --- |
|  | Mean | Std | Min | Max | Skewness | Kurtosis | Absence % | Presence % |
| Jaundice | 0.200334 | 0.40027 | 0 | 1 | 1.497391 | 0.242181 | 79.96658 | 20.03342 |
| Abdominal_D<br>iscomfort | 0.295291 | 0.456196 | 0 | 1 | 0.897503 | -1.19449 | 70.47085 | 29.52915 |
| Back_Pain | 0.250664 | 0.433416 | 0 | 1 | 1.15062 | -0.67607 | 74.93365 | 25.06635 |
| Weight_Loss | 0.34739 | 0.476164 | 0 | 1 | 0.641028 | -1.58908 | 65.26098 | 34.73902 |

| **Stage IV (n = 19918)** |  |  |  |  |  |  |  |  |
| --- | --- | --- | --- | --- | --- | --- | --- | --- |
|  | Mean | Std | Min | Max | Skewness | Kurtosis | Absence % | Presence % |
| Jaundice | 0.196305 | 0.397212 | 0 | 1 | 1.529173 | 0.338371 | 80.36952 | 19.63048 |
| Abdominal_D<br>iscomfort | 0.298474 | 0.4576 | 0 | 1 | 0.880819 | -1.22416 | 70.15263 | 29.84737 |
| Back_Pain | 0.252234 | 0.434306 | 0 | 1 | 1.141003 | -0.69811 | 74.77658 | 25.22342 |
| Weight_Loss | 0.351742 | 0.477526 | 0 | 1 | 0.620957 | -1.61441 | 64.82579 | 35.17421 |

**Supplementary Table 2.** Summary of centrality indices by stage.

| **Stage I (n = 4937)** |  |  |  |
| --- | --- | --- | --- |
|  | Strength | Betweenness | Closeness |
| Jaundice | 0.4370 | 0.0000 | 6.8655 |
| Abdominal_Discomfort | 0.4020 | 0.3333 | 7.4631 |
| Back_Pain | 0.2244 | 0.0000 | 7.0948 |
| Weight_Loss | 0.2075 | 0.0000 | 7.3897 |

| **Stage II (n = 10173)** |  |  |  |
| --- | --- | --- | --- |
|  | Strength | Betweenness | Closeness |
| Weight_Loss | 0.2679 | 0.6667 | 12.2686 |
| Jaundice | 0.2212 | 0.0000 | 7.7685 |
| Abdominal_Discomfort | 0.1270 | 0.6667 | 12.2686 |
| Back_Pain | 0.0614 | 0.0000 | 8.1660 |

| **Stage III (n = 14968)** |
| --- |

|  | Strength | Betweenness | Closeness |
| --- | --- | --- | --- |
| Jaundice | 0.1730 | 0.3333 | 7.7084 |
| Back_Pain | 0.1159 | 0.0000 | 4.6156 |
| Weight_Loss | 0.0571 | 0.0000 | 5.7962 |
| Abdominal_Discomfort | 0.0000 | 0.0000 | 0.0000 |

| **Stage IV (n = 19918)** |  |  |  |
| --- | --- | --- | --- |
|  | Strength | Betweenness | Closeness |
| Jaundice | 0.1508 | 0.3333 | 8.8418 |
| Abdominal_Discomfort | 0.0788 | 0.0000 | 5.8068 |
| Weight_Loss | 0.0720 | 0.0000 | 5.9850 |
| Back_Pain | 0.0000 | 0.0000 | 0.0000 |

**Supplementary Table 3.**
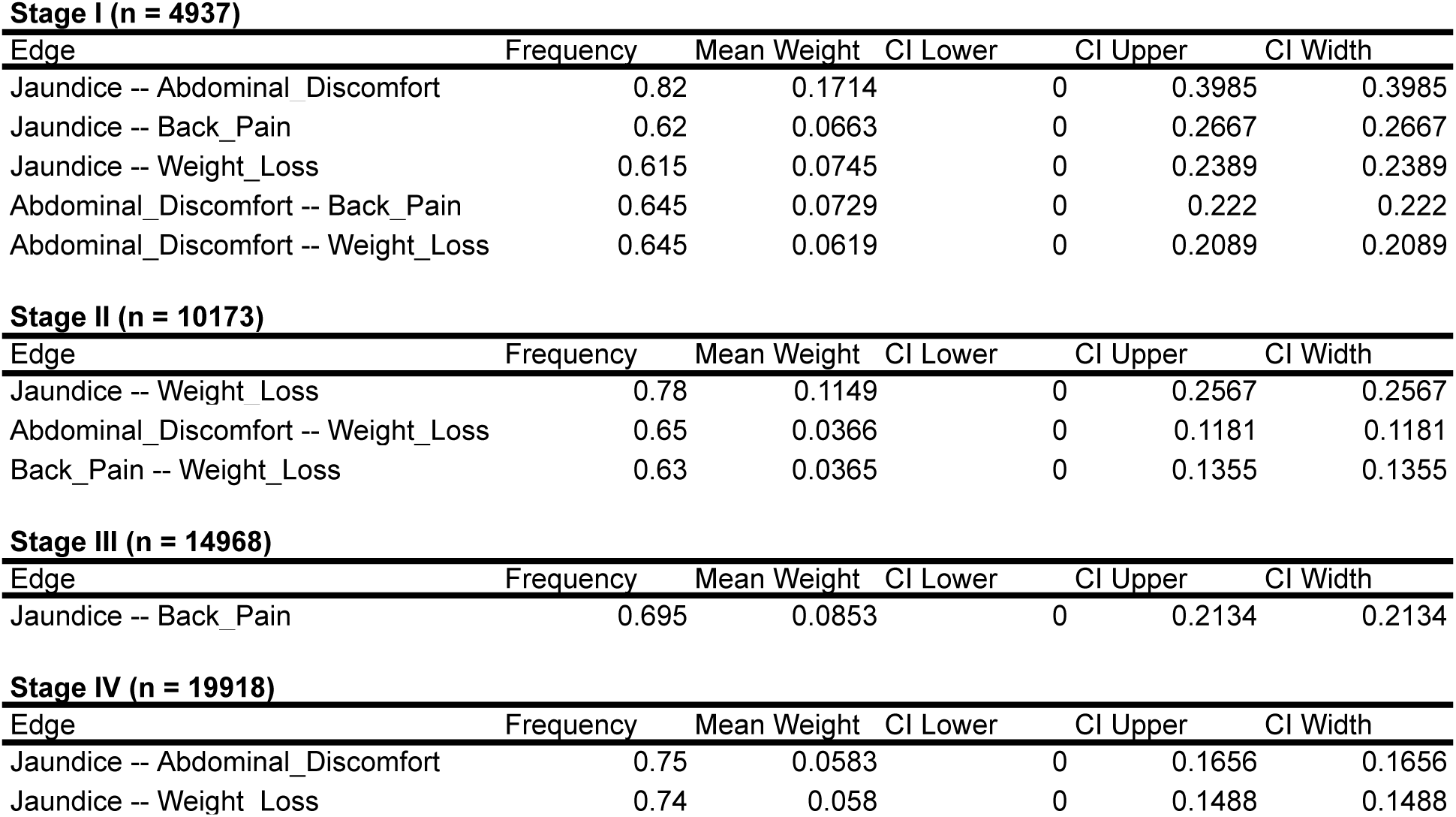
Summary of stable edges with 95% confidence intervals.

**Supplementary Table 4.**
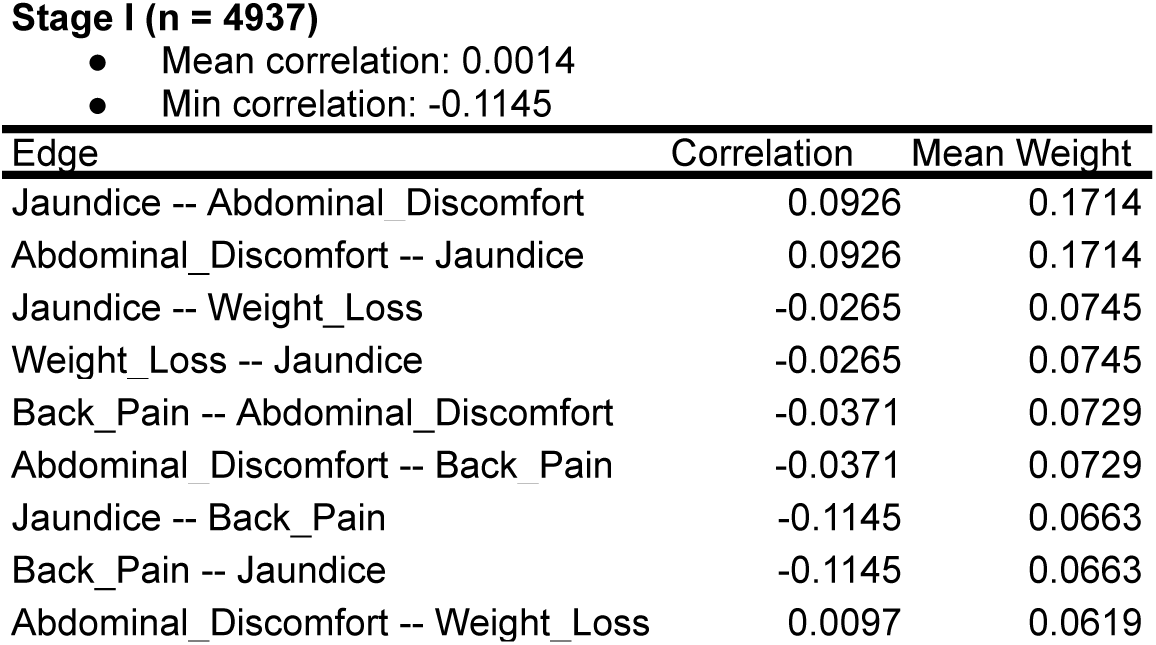
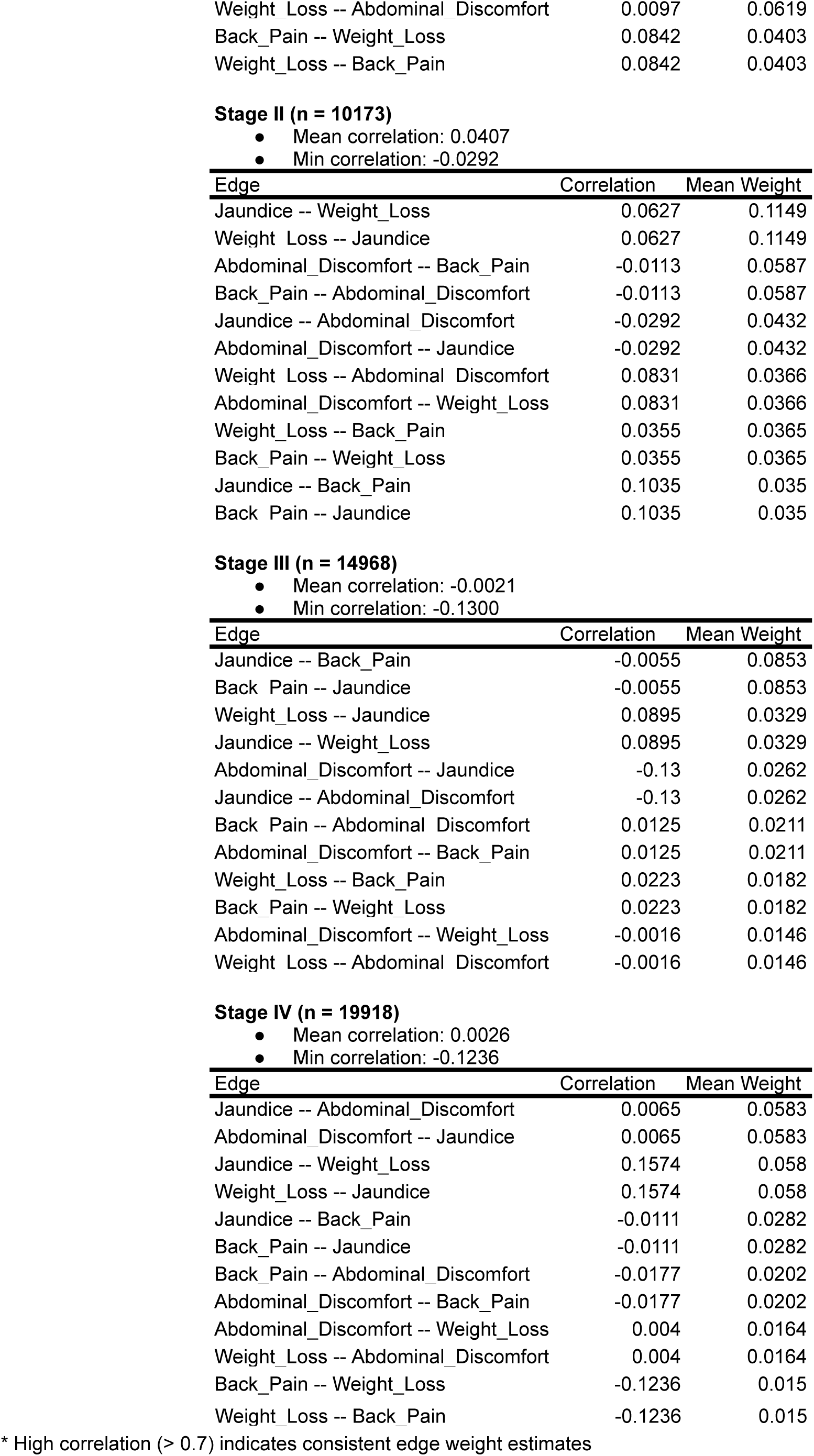
Summary of bootstrap consistency - edge weight correlations between first and second half of bootstrap samples (by drop percentage)

**Supplementary Data 5.**
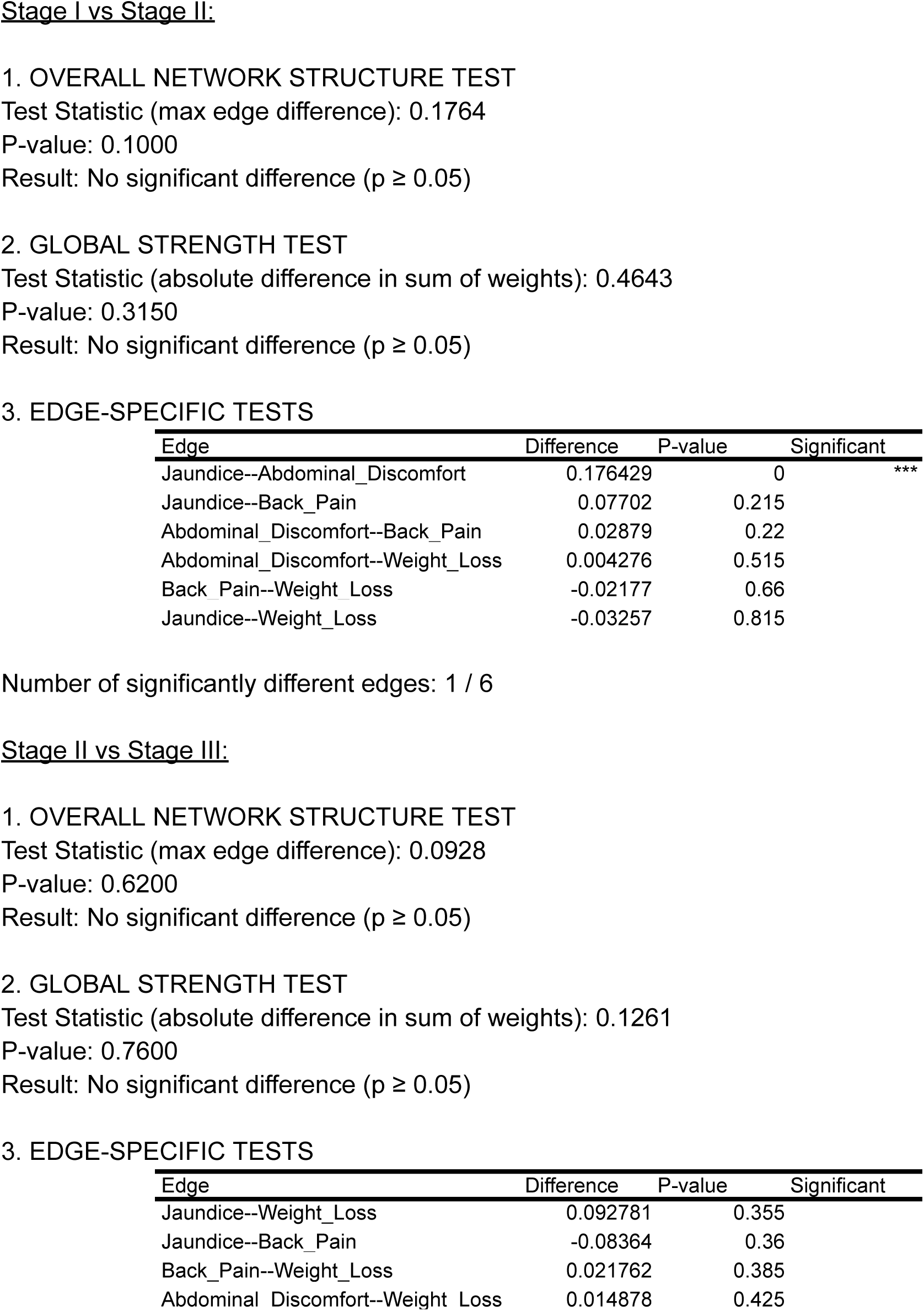
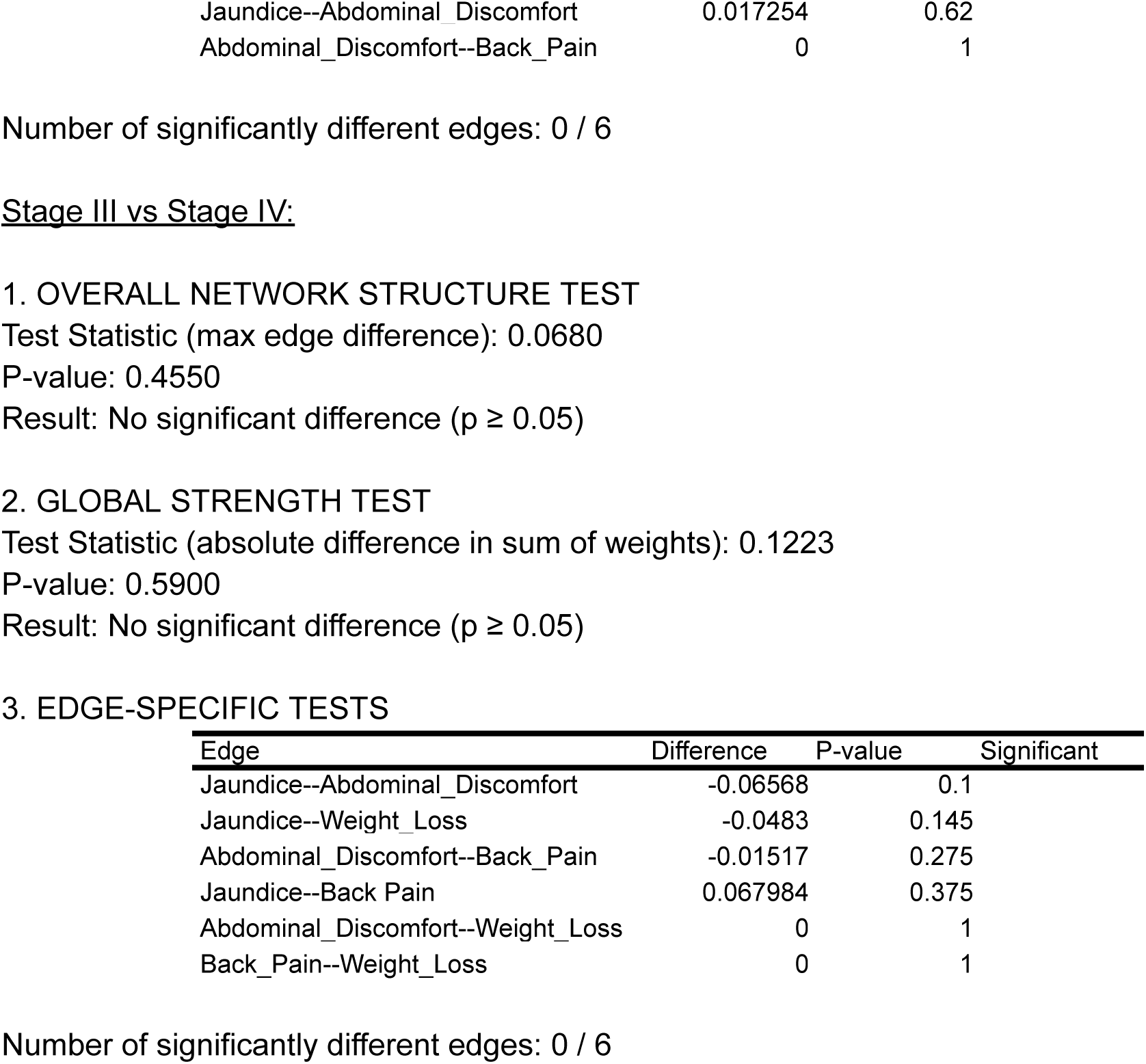
Network comparison test results.

3. EDGE-SPECIFIC TESTS
| Edge | Difference | P-value | Significant |
| --- | --- | --- | --- |
| Jaundice--Abdominal_Discomfort | 0.176429 | 0 | *** |
| Jaundice--Back_Pain | 0.07702 | 0.215 |  |
| Abdominal_Discomfort--Back_Pain | 0.02879 | 0.22 |  |
| Abdominal_Discomfort--Weight_Loss | 0.004276 | 0.515 |  |
| Back_Pain--Weight_Loss | -0.02177 | 0.66 |  |
| Jaundice--Weight_Loss | -0.03257 | 0.815 |  |

3. EDGE-SPECIFIC TESTS
| Edge | Difference | P-value | Significant |
| --- | --- | --- | --- |
| Jaundice--Weight_Loss | 0.092781 | 0.355 |  |
| Jaundice--Back_Pain | -0.08364 | 0.36 |  |
| Back_Pain--Weight_Loss | 0.021762 | 0.385 |  |
| Abdominal_Discomfort--Weight_Loss | 0.014878 | 0.425 |  |

|  |  |  |
| --- | --- | --- |
| Jaundice--Abdominal Discomfort | 0.017254 | 0.62 |
| Abdominal_Discomfort--Back_Pain | 0 | 1 |

3. EDGE-SPECIFIC TESTS
| Edge | Difference | P-value | Significant |
| --- | --- | --- | --- |
| Jaundice--Abdominal_Discomfort | -0.06568 | 0.1 |  |
| Jaundice--Weight_Loss | -0.0483 | 0.145 |  |
| Abdominal_Discomfort--Back_Pain | -0.01517 | 0.275 |  |
| Jaundice--Back Pain | 0.067984 | 0.375 |  |
| Abdominal_Discomfort--Weight_Loss | 0 | 1 |  |
| Back_Pain--Weight_Loss | 0 | 1 |  |

Number of significantly different edges: 0 / 6

**Supplementary Table 6.** Summary of details in difference in network edges.

Stage I → Stage II:
| Edge | Difference | Direction |
| --- | --- | --- |
| Jaundice -- Abdominal_Discomfort | -0.1327 | Stronger in earlier stage |
| Jaundice -- Back_Pain | -0.05988 | Stronger in earlier stage |
| Abdominal_Discomfort -- Weight_Loss | -0.04252 | Stronger in earlier stage |
| Jaundice -- Weight_Loss | 0.041512 | Stronger in later stage |
| Back_Pain -- Weight_Loss | -0.02262 | Stronger in earlier stage |
| Abdominal_Discomfort -- Back_Pain | -0.01019 | Stronger in earlier stage |

Stage II → Stage III:
| Edge | Difference | Direction |
| --- | --- | --- |
| Jaundice -- Weight_Loss | -0.0933 | Stronger in earlier stage |
| Jaundice -- Back_Pain | 0.051194 | Stronger in later stage |
| Jaundice -- Abdominal_Discomfort | -0.0245 | Stronger in earlier stage |
| Back_Pain -- Weight_Loss | -0.01948 | Stronger in earlier stage |

Total edges with differences: 4 Stage III → Stage IV:
| Edge | Difference | Direction |
| --- | --- | --- |
| Jaundice -- Back_Pain | -0.0664 | Stronger in earlier stage |
| Jaundice -- Abdominal_Discomfort | 0.032494 | Stronger in later stage |
| Jaundice -- Weight_Loss | 0.014916 | Stronger in later stage |

Total edges with differences: 3

